# Life’s Essential 8 and Incident Cardiovascular Disease: Validation Using Real World Data from Consumer Devices in the All of Us Research Program

**DOI:** 10.64898/2026.05.07.26352702

**Authors:** Julien O. Tremblay, Jeffrey Annis, Hiral Master, Adnan Cakar, Peyton Coleman, Kelsie Full, Douglas M Ruderfer, Tali Elfassy, Evan Brittain

**Affiliations:** Division of General Internal Medicine, Vanderbilt University Medical Center, Nashville, TN, USA; Vanderbilt Institute for Clinical and Translational Research, Vanderbilt University Medical Center, Nashville, TN, USA; Division of Cardiovascular Medicine, Department of Medicine, Vanderbilt University Medical Center, Nashville, TN; Department of Health Sciences, School of Medicine, University of North Carolina at Chapel Hill, Chapel Hill, NC, USA; Vanderbilt Genetics Institute, Vanderbilt University Medical Center, Nashville TN; Division of Nephrology and Hypertension, University of Miami, Miami, FL, USA

## Abstract

**Background:** The Life’s Essential 8 (LE8) metric is a well-validated tool to assess cardiovascular health. The tool relies on self-reported physical activity (PA) and sleep data which may be subject to recall bias when compared with objective device-derived data. We used objectively captured device data from Fitbit devices linked to the electronic health record (EHR) from the All of Us Research Program (AoURP) to examine the association between LE8 and incident cardiovascular disease (CVD).

**Methods:** We analyzed AoURP participants with ≥6 months of Fitbit-derived PA and sleep data from 2009 to 2023. Remaining LE8 components were obtained via EHR and combined with Fitbit components to calculate LE8 scores. Cox proportional hazards models analyzed the association between LE8 scores and a composite CVD outcome (myocardial infarction, coronary artery disease, heart failure, stroke, and peripheral artery disease). Relative explained variation (REV) assessed the contribution of each LE8 component to model performance. We modeled the impact of plausible changes in weekly activity and sleep on the composite CVD outcome.

**Results:** 11,542 participants were included (50.1 years [IQR: 35.9, 61.7], 74% female, 81% white) with a median monitoring duration of 4.48 years [2.00, 6.87]. The median LE8 score was 68.1 [60.6, 74.4]. Higher LE8 score was linearly associated with lower CVD risk (HR = 0.74; CI, 0.69-0.80) per 10-point increase. Risk of MI, CAD, HF, PAD, and stroke showed similar independent associations with LE8 scores. Among LE8 components, physical activity had the highest median REV 0.35 [0.21, 0.47], followed by blood pressure (0.23, CI = 0.11-0.36) and blood glucose (0.14, CI = 0.05-0.24). Increasing weekly moderate to vigorous physical activity by 30 minutes (120min to 150min) decreased the risk of incident CVD by 23% (HR=0.77; CI, 0.721-0.81), and increasing sleep duration from 4-5 hours to 7-9 hours decreased the risk of incident CVD by 35% (HR=0.65; CI, 0.50-0.84).

**Conclusion:** These results underscore the potential of calculating the LE8 score using objective PA and sleep data from consumer devices and highlight the disproportionate impact of lifestyle behaviors on CVD risk among patients seeking care. Consumer wearable devices offer valuable information when included in cardiovascular risk assessment.

## Introduction

Established by the American Heart Association in 2010, Life’s Simple 7 (LS7) was formulated to define and promote cardiovascular health (CVH) at the individual and population level^1^. Defined by health behaviors and factors, LS7 allowed practitioners to easily quantify cardiovascular risk profiles and patients to strive for optimal CVH^1^. Ideal LS7 was subsequently associated with lower risk of cardiovascular disease (CVD), dementia, all-cause mortality, and a multitude of other chronic conditions^2–5^. The AHA announced an updated construct in 2022, Life’s Essential 8 (LE8), that included an updated scoring system with the addition of sleep health^6^.

While many studies have confirmed the applicability of the metric to predict CVD and other related conditions^7–12^, a possible limitation of these studies is the subjective measurement of some health behaviors. For example, activity levels and sleep duration in LE8 rely on self-report, which is susceptible to recall and social desirability bias and may not capture habitual behavior over time ^13–19^, weakening the discerning nature of the LE8 metric. Published studies using objective markers (actigraphy, accelerometry) to calculate the LE8 score are limited to short time periods (</= 7 days) that may not represent habitual behavior ^20,21^. Indeed, the presidential advisory group that developed the LE8 metrics recognized these limitations and called for data incorporating consumer wearable devices to capture long-term quantitative behaviors ^6^.

In response to this call from the AHA Presidential Advisory, we evaluate the LE8 score in participants in the All of Use Research Program. The AoURP is a large NIH-funded precision medicine initiative that includes individuals with electronic health record (EHR) data linked to long-term, quantitative behavior monitoring using Fitbit devices. We thus examined the association between LE8 score and incident cardiovascular disease in patients real world data from the EHR and consumer wearable devices.

## Methods Participants

Participants aged over 18 years were enrolled after informed consent at clinics and medical centers across the country that comprise the AoURP network. A detailed description of the AoURP has been previously published^22^. The All of Us protocol and materials have been approved by the All of Us Institutional Review Board. For the present study, we used the AoURP Registered Tier Dataset version 8 (R2024Q3R4) available on the AoURP Research Workbench, a secure cloud-based platform. This dataset includes patient specific information on physical measures, surveys, and wearable device data, specifically Fitbit data, and is linked to the EHR. Participants who were included in this study were enrolled from May 31 2017 to October 1, 2023. Our analyses utilized participants who had linked Fitbit data for at least 6 months and agreed to share their EHR data. All data are publicly available at www.allofus.nih.gov following proposal submission and approval.

## Study Cohort

Participants provided wearable device data in one of two ways. Most participants contributed Fitbit data at the time of enrollment from devices they already owned [Bring Your Own Device program (BYOD)]. Alternatively, some AoURP participants (who identified with one or more communities underrepresented in biomedical research) were provided Fitbit devices at no cost by the program as part of the *Wearables Enhancing All of Us Research* (WEAR) study^23,24^. We included All of Us participants who were >18 years of age, had at least 6 months of Fitbit data with valid day wear (>10 hours/day) and valid sleep data, and agreed to share their EHR data. We excluded participants with baseline prevalent CVD (as defined below) (**Figure S1**). The median number of missing LE8 categories was 1 IQR [0, 3], and sensitivity analyses were preformed to exam the impact of missing components on overall score performance.

## LE8 Components

Information on cardiovascular health metrics, except for diet, were obtained from the EHR, including body mass index, blood lipids, blood glucose control, and blood pressure, and from survey data on nicotine exposure. Measurements could be drawn from any time prior to the start date, whereas post–start date measurements were restricted to those occurring within the subsequent 180 days.

The blood glucose LE8 component was calculated using HbA1c and diabetes status, where diabetes status was defined using the definition from the All of Us phenotype library^25,26^. Nicotine exposure was derived from self-reported smoking history and current smoking behavior from the All of Us Lifestyle survey. For the blood pressure component, categories were assigned based on systolic and diastolic values, as well as antihypertensive status (subtracting 20 points when prescribed). For the blood lipids and BMI components, we used non-HDL-C and BMI reported in the EHR, respectively. For each participant, the EHR measurement closest to the start of the Fitbit monitoring period was used. Using methodology described in detail by Lloyd-Jones et al, we defined each cardiovascular health metric on a scale of 0 to 100^6^. LE8 score was subsequently calculated as an average of all 7 available scaled health metrics, diet excluded.

## Fitbit Data

*All of Us* Fitbit data include daily summary variables for PA (sedentary minutes, lightly active minutes, moderately active minutes, and very active minutes) and sleep (duration and stages). According to the Fitbit Web API Data dictionary, metabolic equivalent of task (MET) definitions for Fitbit PA summary variables are: <1.5 METs, sedentary minutes; 1.5–3.0 METs, lightly active minutes; 3.0–6.0 METS, fairly (or moderately) active minutes; >6.0 METs, very (or vigorously) active minutes. Based on NHANES accelerometry research and All of Us study using Fitbit data, a valid day was considered at least 10 hours of wear time with at least 100 steps reported during that time^27–30^. Similar assessments of Fitbit derived sleep data have shown that removing days where no heart rate data is available, >24 hours of sleep data are logged, or <0 minutes of sleep data are logged improves the accuracy and reproducibility of the overall study^27^. Daily sleep metrics were retained only if total recorded sleep duration was greater than 0 minutes and did not exceed 24 hours. Participants were required to have no more than 30% of observed days with sleep duration exceeding 4 hours^31^. Analyses were restricted to adult participants aged 18 years or older. Only sleep episodes designated by Fitbit as “main sleep” were included, excluding naps and secondary sleep segments.

For Fitbit metrics, we took the average over the monitoring period. Follow-up data were restricted to observations occurring before each participant’s event date. To reduce reverse causation and ensure adequate exposure ascertainment, a 180-day blanking period was enforced. Specifically, participants were required to remain event-free for at least 180 days following the start of observation; individuals whose event date occurred within the first 180 days were excluded. Dietary information is not available in the AoURP. To address this limitation, we provide range estimates within our hazard ratio plots that model either the best or worst possible diet score for every individual. To quantify the physical activity component of LE8, we calculated time spent in moderate and vigorous activity minutes per week by adding Fitbit measured time spent in fairly active and very active minutes. Sleep duration was calculated using daily mean sleep time as reported by the device.

## Outcomes

We defined incident CVD as a composite of incident myocardial infarction (MI), coronary artery disease (CAD), heart failure (HF), peripheral artery disease (PAD) or stroke (both ischemic and hemorrhagic). We derived a composite outcome by selecting the first event date of the component disease listed above. All definitions were based on ICD9/10 codes (**Table S1**)^32,33^.

## Statistical Analysis

Unadjusted and sex and age adjusted Cox proportional hazards model were used to estimate the association between baseline LE8 score and time to first incident composite CVD event and with each its component separately. Additional models were fit with LE8 sub scores as predictors to evaluate their separate associations with incident composite CVD. For continuous values, hazard ratios were estimated per 10-unit increase in LE8 score or correspond to contrasts evaluated at prespecified values. The proportional hazards assumption was assessed using Schoenfeld residuals. The baseline LE8 score was anchored to the date of the first available valid Fitbit monitoring record. Participants were censored at their last medical encounter, defined as the most recent laboratory or vital sign measurement, diagnosis, procedure, or prescription. Fitbit-derived physical activity and sleep metrics were averaged across the interval from baseline to last follow-up. Other LE8 components were assigned using the closest measurement prior to baseline or within 180 days afterward. Missing elements of the LE8 score (blood pressure, blood lipids, BMI, nicotine exposure) were imputed using multiple imputation with bootstrap resampling and predictive mean matching (10 imputations)^34^. Final LE8 scores were then calculated incorporating imputed values. Sensitivity analyses were conducted with the constraint that the maximum number of imputed elements remained at or below a specified threshold (1–3 elements). The relative importance of each component was assessed with relative explained variation (REV), the proportion of variation due to each predictor. REV medians and 95% confidence intervals were based on 300 bootstrap samples. Group comparisons for continuous variables were performed using the Wilcoxon rank-sum test or Kruskal-Wallace test for >2 groups, and categorical variables were compared using the Pearson chi-square test. A two-*sided p*-value < 0.05 was considered statistically significant. All analyses were conducted in R version 4.5 (R Project, https://www.r-project.org) on the All of Us Researcher Workbench.

### Exploratory Analyses

Phenome-wide association study: Clinical phenotypes were defined using ICD-9 and ICD-10 codes mapped to phecodes using a standard phecode mapping system^35^. For each participant and phecode, the earliest recorded diagnosis date was extracted. Incident phenotypes were defined as diagnoses occurring more than 180 days after baseline. Diagnoses within or before the first 180 days of monitoring were excluded from the sample.

A phenome-wide association study was conducted by fitting separate logistic regression models for each phecode, with case–control status as the dependent variable. The primary independent variable was the CVH score per 10-point increase. All models were adjusted for age at baseline, sex, and calendar time (baseline date). Odds ratios and 95% confidence intervals were estimated for each phecode.

Models were fit independently for each phenotype using complete-case data. Multiple testing was addressed using Bonferroni correction based on the total number of phecodes tested. Associations meeting the Bonferroni-adjusted significance threshold were considered statistically significant.

Psychosocial determinants of health: Social and neighborhood determinants of health were assessed using validated self-report survey instruments. Perceived neighborhood walkability and bicycling infrastructure were measured using four items from the Physical Activity Neighborhood Environment Survey (PANES), assessing access to recreational facilities, pedestrian and bicycle infrastructure, proximity to destinations, and public transit, where lower scores indicate poorer neighborhood accessibility and higher scores indicate greater accessibility. Loneliness was assessed using the 8-item UCLA Loneliness Scale, capturing subjective feelings of social isolation and disconnectedness^36^, where lower scores indicate lower perceived loneliness and higher scores indicate greater loneliness. Everyday discrimination was measured using nine items assessing the frequency of routine unfair treatment^37,38^, with lower scores reflecting less frequent experiences of discrimination and higher scores reflecting more frequent discrimination. Perceived social support was assessed with eight items from the RAND Medical Outcomes Study Social Support Survey, capturing emotional, informational, tangible, and affectionate support^39^, where lower scores indicate lower perceived social support and higher scores indicate greater perceived social support. Perceived stress was measured using the 10-item Perceived Stress Scale, assessing the degree to which life situations were experienced as stressful^40^, with lower scores indicating lower perceived stress and higher scores indicating greater perceived stress.

All items were rated on Likert-type frequency or agreement scales, scored according to published guidelines, and reverse-coded as appropriate so that higher values reflected greater levels of the underlying construct. For each measure, participant-level scores were calculated as the mean of available item responses, allowing for partial completion. All instruments have demonstrated reliability and validity in prior population-based research.

## Results

### Population Characteristics

We identified 11,542 AoURP participants with long-term Fitbit data connected to their EHR to include in analysis (**Figure S1**). Among these, 19% of participants were enrolled through WEAR and 81% through BYOD. Median age in the population was 50.1 IQR [35.9.0, 61.7] including 74% women and 81% white, non-Hispanic/Latino adults. Overall, 70% had completed college education and 42% had an income >$100,000 per year (**Table 1**). Overall LE8 score with assumed median diet was 70.2 IQR [62.7, 76.4]. Modeling ideal and worst diet scores changed the overall score to 70.9 IQR [63.4, 77.2] and 69.4 IQR [61.9, 75.6], respectively. **Figure 1** displays the frequency and distribution LE8 component scores in the study population.

**Figure 1:**
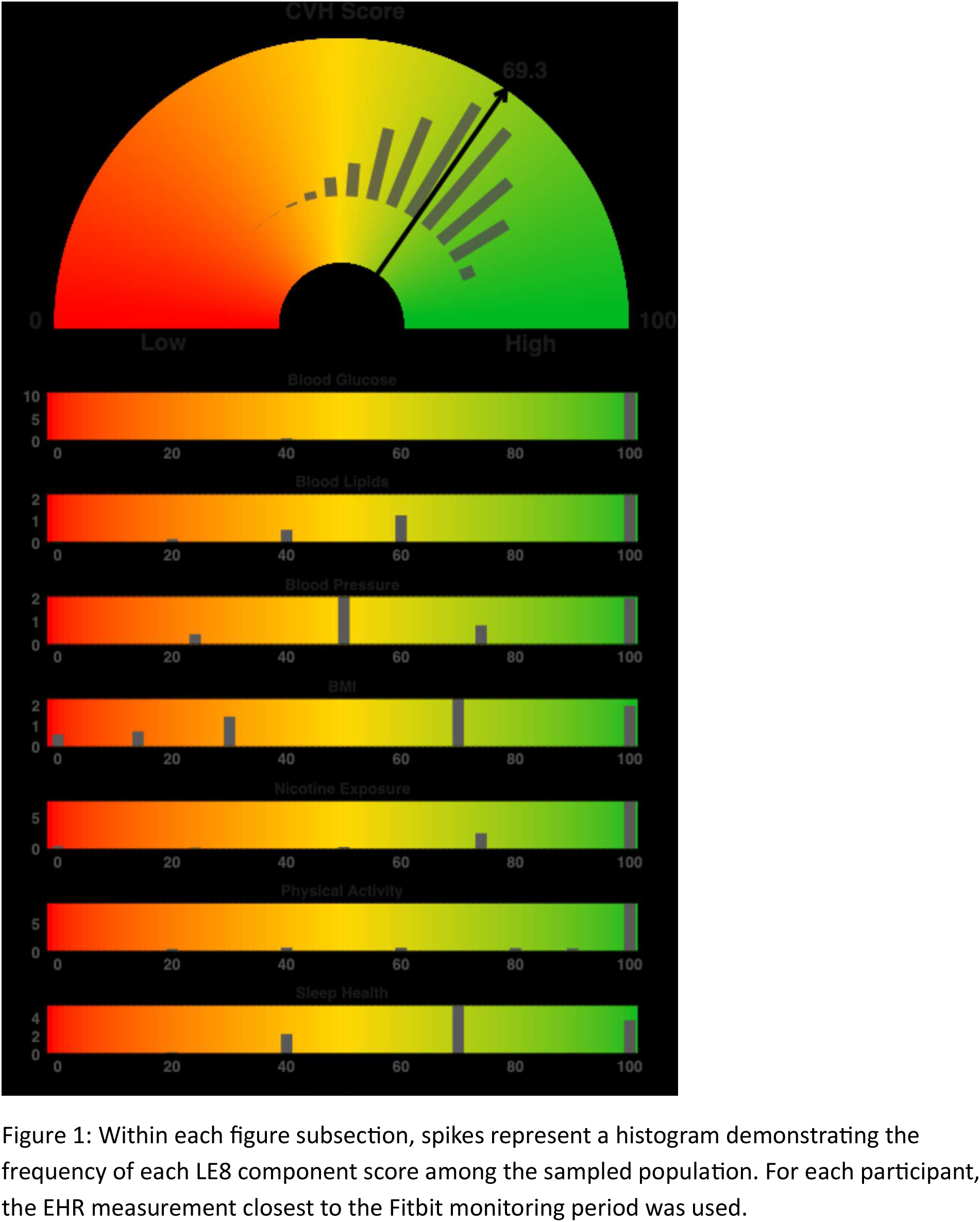
Distribution of LE8 Scores Amongst the Sampled Population

**Table 1.**
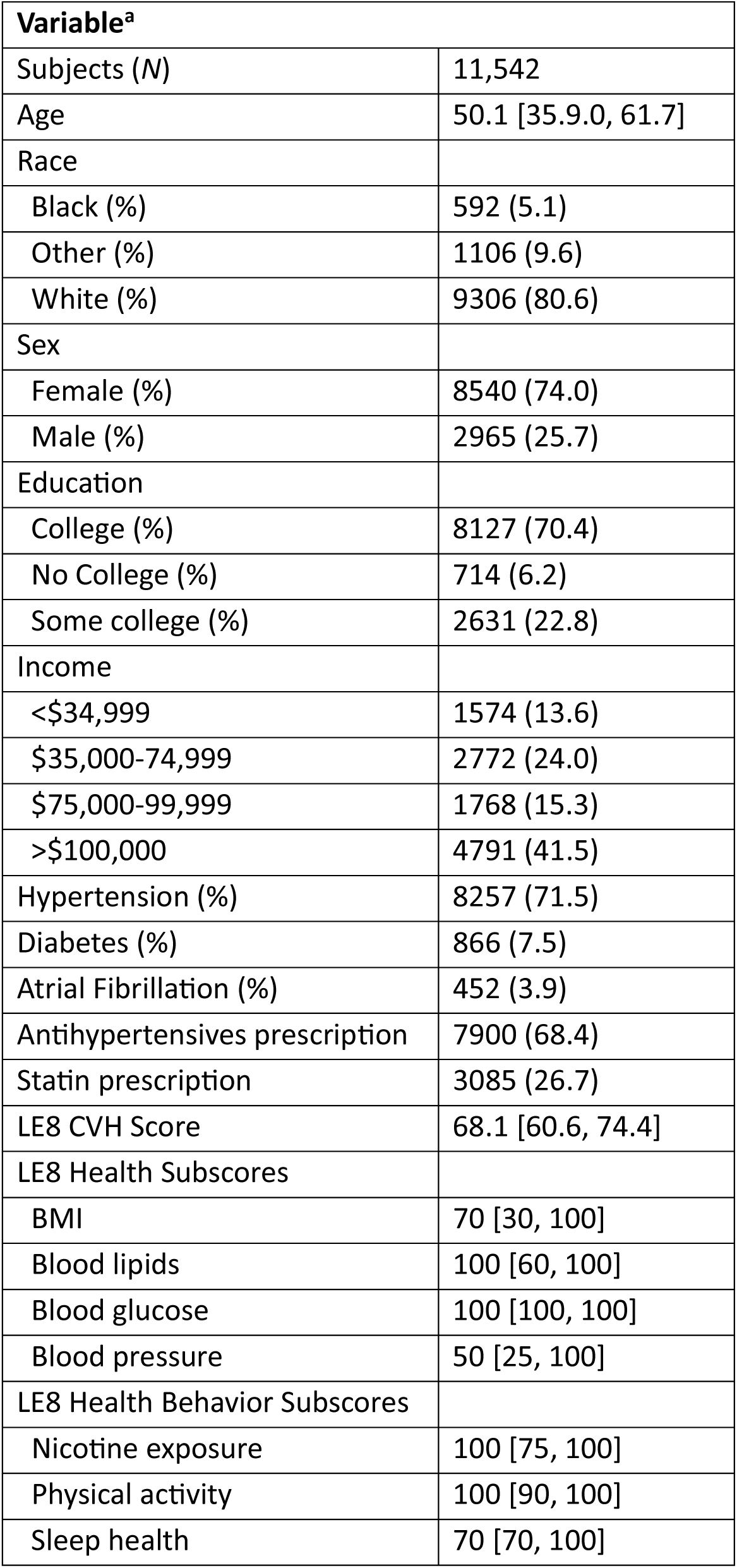
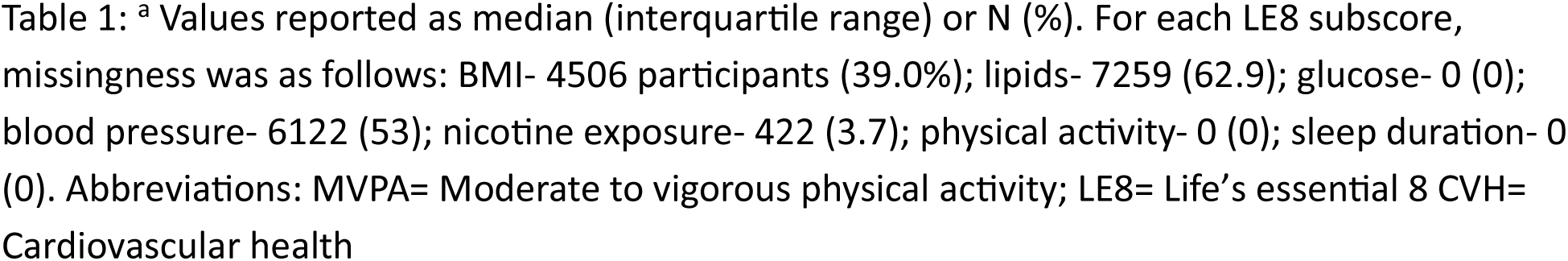
Patient characteristics.

<sup>a</sup> Values reported as median (interquartile range) or N (%).
| **Variable<sup>a</sup>** |  |
| --- | --- |
| Subjects (N) | 11,542 |
| Age | 50.1 [35.9.0, 61.7] |
| Race |  |
| Black (%) | 592 (5.1) |
| Other (%) | 1106 (9.6) |
| White (%) | 9306 (80.6) |
| Sex |  |
| Female (%) | 8540 (74.0) |
| Male (%) | 2965 (25.7) |
| Education |  |
| College (%) | 8127 (70.4) |
| No College (%) | 714 (6.2) |
| Some college (%) | 2631 (22.8) |
| Income |  |
| <\$34,999 | 1574 (13.6) |
| \$35,000-74,999 | 2772 (24.0) |
| \$75,000-99,999 | 1768 (15.3) |
| >\$100,000 | 4791 (41.5) |
| Hypertension (%) | 8257 (71.5) |
| Diabetes (%) | 866 (7.5) |
| Atrial Fibrillation (%) | 452 (3.9) |
| Antihypertensives prescription | 7900 (68.4) |
| Statin prescription | 3085 (26.7) |
| LE8 CVH Score | 68.1 [60.6, 74.4] |
| LE8 Health Subscores |  |
| BMI | 70 [30, 100] |
| Blood lipids | 100 [60, 100] |
| Blood glucose | 100 [100, 100] |
| Blood pressure | 50 [25, 100] |
| LE8 Health Behavior Subscores |  |
| Nicotine exposure | 100 [75, 100] |
| Physical activity | 100 [90, 100] |
| Sleep health | 70 [70, 100] |

### Association of LE8 Metrics and Incident Cardiovascular Disease Composite Outcome

Participants were followed for a median of 4.48 years IQR [2.00, 6.87] from baseline to censoring or event. During follow-up, there were 578 incident myocardial infarctions, 330 coronary artery disease events, 169 cases of heart failure, 133 cases of peripheral artery disease, and 126 strokes. We derived a composite outcome by selecting the first event date of the component disease listed above, which resulted in 769 incident events. A sex and age adjusted Cox proportional hazards model revealed LE8 scores were significantly and inversely associated with incident CVD, *p <* 0.001 (**Figure 2**). A 10-point increase in the LE8 score was associated with a 26% decreased risk of CVD (HR = 0.74, 95% CI = 0.68-0.81).

**Figure 2:**
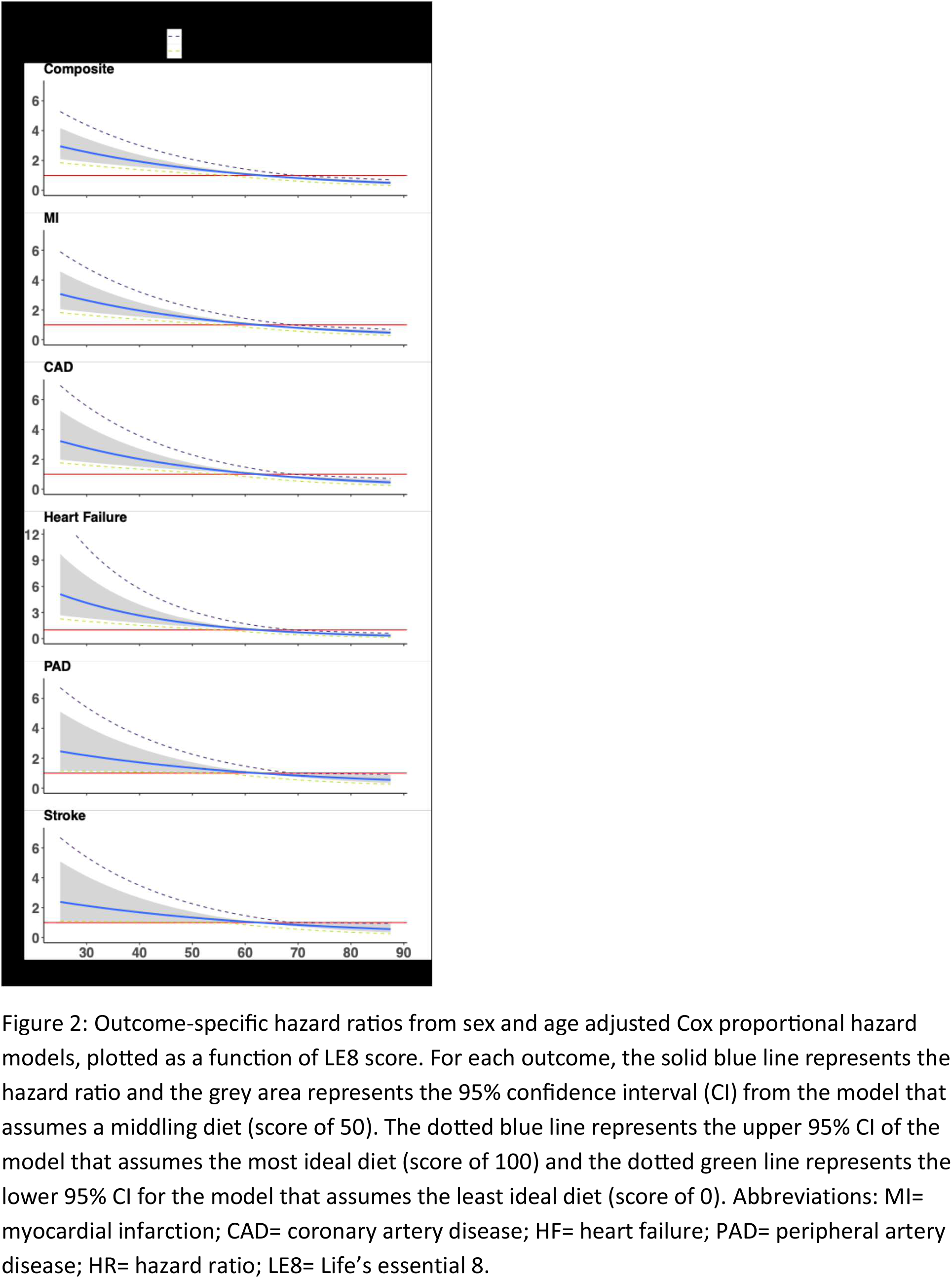
Outcome Associated Hazard Ratios as a Function of LE8

We next modeled incident CVD risk adjusting for all components of the LE8 score. The hazard of composite CVD event was independently associated with physical activity, blood glucose, sleep duration, blood pressure, and nicotine exposure. Physical activity had the highest median REV 0.35 [0.21, 0.47], followed by blood pressure (0.23, CI = 0.11-0.36) and blood glucose (0.14, CI = 0.05-0.24) (**Figure 3**). Chi square analyses demonstrated similar findings.

**Figure 3:**
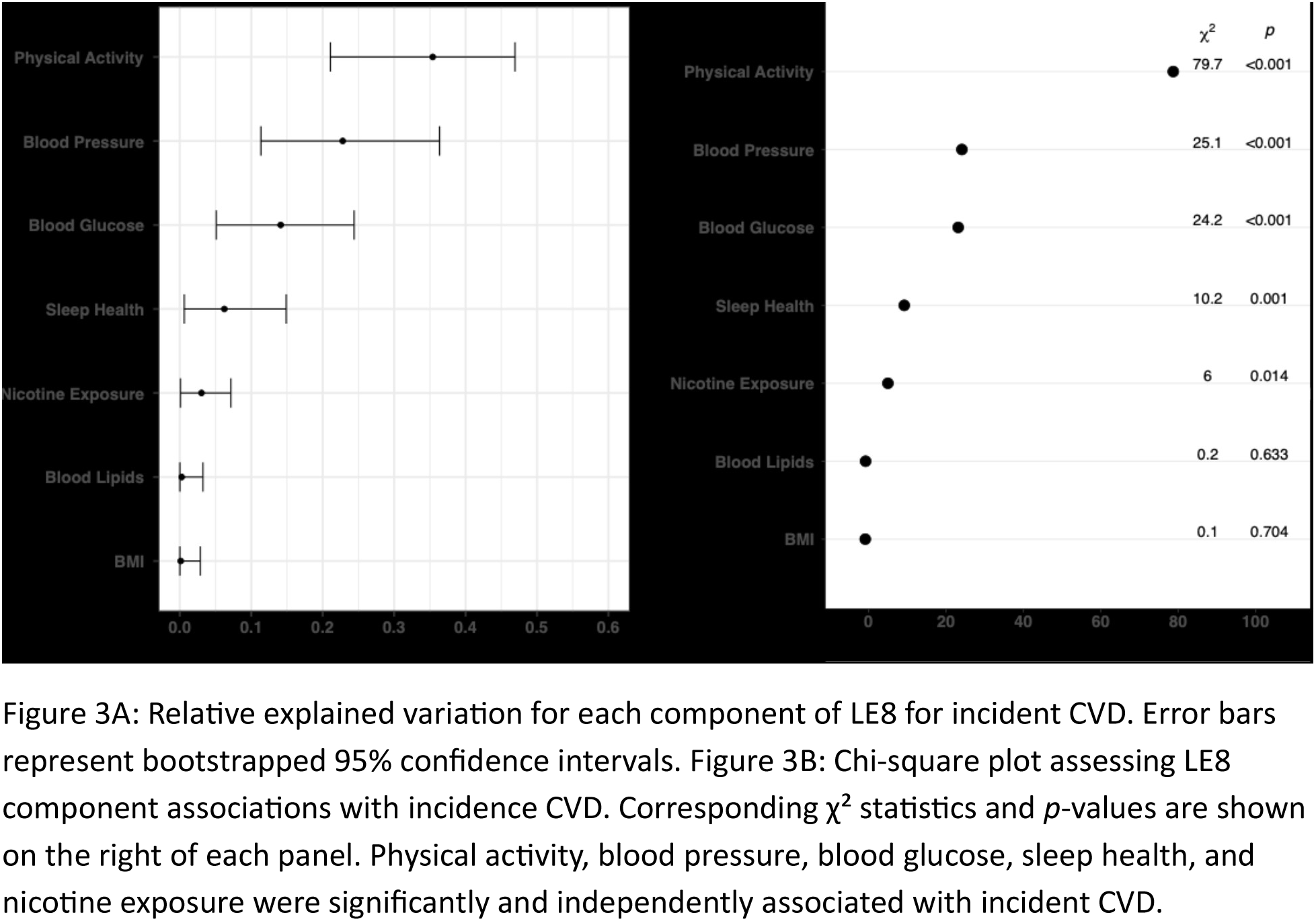
Relative explained variation plot for each predictor

Sensitivity analyses were performed to evaluate the impact of imputation by restricting analyses to participants with no more than 1, 2, or 3 missing LE8 components (**Table S2**). Across these thresholds, Cox proportional hazards models produced coefficients and *p*-values for the LE8 score that were consistent with the primary analysis. This stability extended to subgroup effects, as no statistically significant interactions were observed between CVH score and age (*p* = 0.53) or sex (*p* = 0.33).

### Incident MI, CAD, Heart Failure, PAD, and Stroke

We next modeled LE8 score and risk of each individual CVD outcome. When evaluating incident MI alone, each 10-point increase in LE8 score was associated with a 26% decreased risk of MI (HR = 0.74; 95% CI = 0.67-0.83) (**Table 2**). Each 10-point increase in LE8 was also associated with a 27% decreased risk of CAD (HR = 0.73; 95% CI = 0.64-0.83), 35% reduced risk of HF (HR = 0.65; 95% CI = 0.55-0.77), 21% reduced risk of PAD (HR = 0.79; 95% CI = 0.65-0.96), and 21% reduced risk of stroke (HR = 0.79; 95% CI = 0.65-0.97). REV plots revealed similar predictor contributions as the model with composite outcomes with sleep health falling to lower explainability for stroke and PAD **(Figure S2a)**. Chi square analyses demonstrated heterogeneous associations between LE8 components and individual CVD outcomes (**Figure S2b**); however, physical activity remained significantly associated with all outcomes.

**Table 2:** Incident CVD risk associated with cumulative LE8 scores.

Unadjusted and sex and age adjusted cox proportional hazards model were used to model the association between 10-point unit increase in LE8 score and first incident composite CVD event.
| Chronic Disease | Number of Events | HR (95% CI) |
| --- | --- | --- |
| Composite |  |  |
| Unadjusted | 769 | 0.72 (0.67, 0.77) |
| Adjusted |  | 0.74 (0.68, 0.81) |
| MI |  |  |
| Unadjusted | 578 | 0.72 (0.65, 0.80) |
| Adjusted |  | 0.74 (0.67, 0.83) |
| CAD |  |  |
| Unadjusted | 330 | 0.71 (0.63, 0.80) |
| Adjusted |  | 0.73 (0.64, 0.83) |
| HF |  |  |
| Unadjusted | 169 | 0.64 (0.54, 0.75) |
| Adjusted |  | 0.65 (0.55, 0.77) |
| PAD |  |  |
| Unadjusted | 133 | 0.76 (0.64, 0.92) |
| Adjusted |  | 0.79 (0.65, 0.96) |
| Stroke |  |  |
| Unadjusted | 126 | 0.77 (0.63, 0.93) |
| Adjusted |  | 0.79 (0.65, 0.97) |

### Associations Between Changes in Physical Activity and Sleep with Incident CVD

We then modeled how changes in physical activity and sleep affect the risk of incident CVD (**Figure 4)**. Increasing physical activity by 30 minutes/week (90-119 minutes to >150) decreased the risk of incident CVD by 23% (HR = 0.77; CI = 0.721-0.81) whereas decreasing physical activity by 30 minutes per week (90-120 minutes to 60-89) increased the risk of incident CVD by 31% (HR = 1.3; CI = 1.2-1.4). Similarly, increasing sleep duration from 4-5 hours to 7-9 hours decreased the risk of incident CVD by 34% (HR = 0.66; CI = 0.52-0.85).

**Figure 4:**
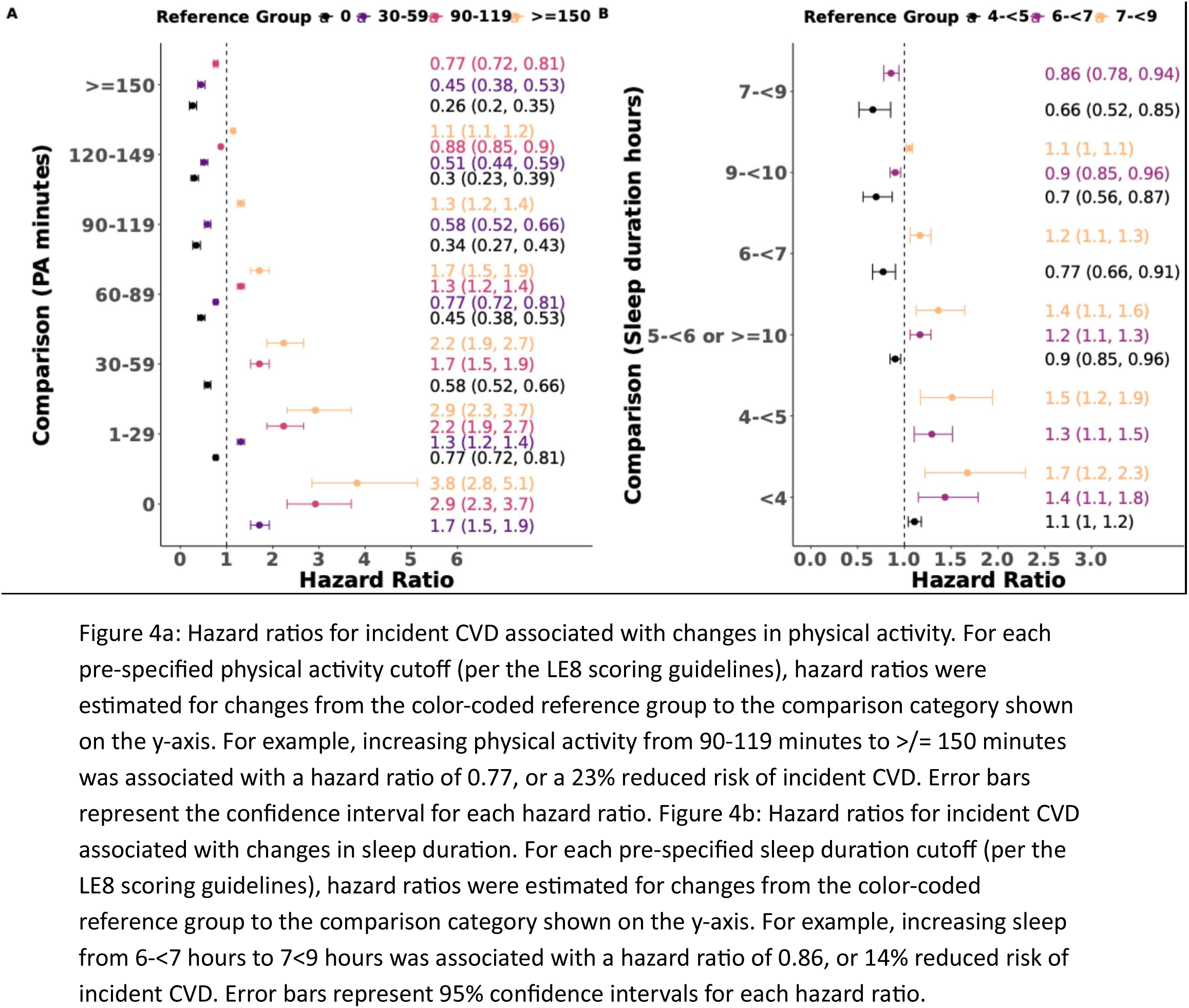
Changes in Physical Activity and Sleep and Risk of Incident CVD

### Exploratory Associations

Because the Presidential Advisory group questioned whether LE8 would be associated with chronic conditions other than CVD, we performed a phenome-wide association study (PheWAS) using LE8 data and incident comorbidities (defined by ICD9/10 codes). Higher LE8 scores were associated with decreased odds of obesity (OR = 0.34, *p* = 7.09 x 10^-22^), type 2 diabetes (OR = 0.44, *p* = 2.44 x 10^-12^), hypertension (OR = 0.56, *p* = 2.35 x 10^-11^), sleep apnea (OR = 0.62, *p* = 5.56 x 10^-10^), and mixed hyperlipidemia (OR = 0.74, *p* = 3.77 x 10^-5^). Higher LE8 scores were also associated with lower odds of incident malabsorptive disorders (OR = 0.72, *p* = 2.41 x 10^-5^), and non-alcoholic liver disease (OR = 0.58, *p* = 2.2 x 10^-8^) (**Figure S3).**

Additionally, we performed several exploratory analyses. First, we evaluated the association between LE8 quartile and psychologic stressors, represented by the UCLA loneliness scale (ULS-8), everyday discrimination scale (EDS), and perceived stress scale (PSS-10) (**Table S3**).

Individuals in Q4 had significantly higher scores compared to Q1 for the physical activity neighborhood environment scale (PANES) (3.0 IQR [2.3, 3.6] vs. 2.75 IQR [2, 3.4], *p* < 0.01) and the Rand social support scale (4.5 IQR [3.9, 5.0] vs. 4.0 IQR [3.1, 4.8], *p* < 0.001).

## Discussion

This study was designed to address knowledge gaps identified by the AHA Presidential Advisory group responsible for developing the LE8 score calling for the validation of the score using real world data sources. We present novel analysis of the AHA’s LE8 score and CVD risk from over 11,000 US adults using long-term, objective behavioral data from consumer wearable devices linked to the EHR. We evaluated the LE8 score using real world data sources, finding an inverse relationship between LE8 score and the risk of incident CVD. We identify a threshold score at which risk begins for the composite and individual outcomes. Importantly, we observed an outsized impact of objectively measured lifestyle behaviors – physical activity and sleep – on CVD outcomes compared with other traditional risk factors. Finally, we present the first use of long-term, objective sleep data in LE8 calculation which strongly corroborated the Advisory group’s decision to include sleep health in the LE8. These findings provide evidence for the clinical value of integrating data from consumer wearables with the EHR for risk assessment using the AHA LE8.

Consistent with prior studies, we found that higher LE8 scores were associated with a decreased risk of incident CVD^41^. When compared with a recent analysis by Ning et al., which assessed an aggregated sample of 6 contemporary cohorts and found a 31% reduced risk of CVD with every 10-point increase in LE8 amongst middle aged adults, we found a 26% reduced risk of incident CVD ^41,42^. In NHANES, investigators found a 29% reduced risk of incident CVD with every 10-point increase in LE8 score^42^. The modest differences in risk reduction in our cohort compared with other modern cohorts likely reflect a combination of factors. Sleep and physical activity were self-reported in prior studies, which may be over- or under-represented on an individual level^13,15,17,43^. Moreover, most participants with Fitbit data in the AoURP owned their devices at the time of enrollment, which may select for a healthier population making risk reduction somewhat harder to achieve compared with other cohort studies.

Our study is novel in examining the impact of objective physical activity and sleep health data from consumer devices on LE8 performance for CVD risk prediction. As expected, sex and age at baseline are the strongest predictors of incident CVD^44,45^. However, when evaluating the relative contribution of individual LE8 categories on model performance, physical activity, blood pressure, blood glucose, and sleep health showed the strongest associations with incident CVD. We demonstrate that increasing physical activity or sleep duration have an important benefit on the risk of incident CVD. This result confirms the primacy of behavioral modification to reduce CVD risk and reaffirms the decades-long emphasis on lifestyle changes as an initial strategy to improve heart health. These findings stand in contrast to a recent analysis by Wadden et al. in U.S. women with breast cancer that found sleep and physical activity were not independently associated with incident CVD when evaluating each of the LE8 components individually. The difference between these findings and those of Wadden et al is likely explained by the difference between self-report and objectively measured behaviors overtime^46^. In fact, one prior study evaluating differences between 7-day accelerometer data and subjective survey data in the calculation of LE8 amongst elderly women found that most participants were reclassified to higher LE8 scores when using accelerometer data compared with survey data^47^. Given our findings in addition to the fact that roughly 1 in 3 of all Americans have a wearable device ^48^ and the widespread availability of EHR data, future studies should prioritize objectively collected data rather than self-report.

Sleep health has emerged as an important determinant of cardiovascular and metabolic disease risk ^20,49–51^. Our finding that sleep duration was robustly associated with incident CVD supports the appropriateness of its inclusion in the Life’s Essential 8 construct. In the most comprehensive prior analysis, Ning et al., using the Lifetime Risk Pooling Project, found that inclusion or exclusion of the sleep metric did not materially change the association between LE8 score and CVD events, a result they attributed to imprecision in self-reported sleep duration^41^. Discrepancies between subjective and objective sleep assessments in research are well documented. Landry et al., found that the Pittsburg Sleep Quality Index (PSQI) classification as either a “good” or a “poor” sleeper lacked predictive validity in determining objective sleep quality^15^. Masaki et al. also found in 2024 that nearly 45% of participants who subjectively reported sufficient sleep were objectively sleep deficient^13^. Sleep health is multidimensional encompassing not only duration but also by continuity, timing, satisfaction, and regularity^51^, and it is further shaped by social determinants that influence cardiovascular risk ^52^ Together, these findings highlight the value of objective, device-derived data for improving the precision and validity of behavioral health metrics central to cardiovascular risk assessment.

As a hypothesis-generating analysis, we further examined the association between objectively measured LE8 metrics and psychological stressors. Participants in the highest (i.e. best) quartile of LE8 scores were more likely to report living in more walkable neighborhoods and having stronger social support compared with those in the lowest quartile. These findings align with prior literature demonstrating that features of the built environment—particularly walkability—are associated with more favorable cardiovascular risk profiles^53,54^. Finally, in a phenome-wide association study (PheWAS), higher LE8 scores were also associated with lower odds of incident major depressive disorder, in addition to expected comorbidities such as sleep apnea and other metabolic conditions. The predecessor construct to LE8, Life’s Simple 7, has similarly been linked to lower risk of incident major depressive disorder^55^. Collectively, these findings underscore the importance of future studies evaluating the relationship between LE8 and chronic diseases other than CVD, especially mental health conditions, particularly given the inclusion of sleep health—an established risk factor for psychiatric illness—within the updated score.

Our study has important strengths that provide valuable insights into the utility of wearable device data in cardiovascular health monitoring at both the individual and population level. As called for by the AHA’s presidential advisory group who developed the LE8 construct, we utilize objectively measured wearable device data in conjunction with linked EHR data to assess cardiovascular risk. While prior studies have used actigraphy or accelerometer data to measure activity, no prior study has used both objective sleep and physical activity data in the same analysis. Moreover, prior studies have been limited by short monitoring periods of days to weeks, whereas the present study benefits from an average of several years of wearable device data. Both prior research ^56^ and the current administration have emphasized wearable device data as an important component to personalize health care^57^, and we present the largest study to date that utilizes objective data sources in the calculation of cardiovascular risk through Life’s Essential 8. Moreover, while previous studies have used EHR data to augment CVD risk prediction^58–60^, we present the first study to assess CVH utilizing LE8 scores using EHR data.

More broadly, our data underscore the importance of physical activity and sleep health in preventing incident CVD. As a relatively recent addition to the AHA’s LE8, sleep health demonstrates a robust association with incident CVD when measured objectively, with effects comparable to other core LE8 metrics.

The findings of this study should be assessed in the context of several limitations. Most importantly, we lacked the ability to incorporate dietary data into our calculation, as the AoURP does not currently incorporate surveys to assess dietary behavior. To mitigate this limitation, we modeled hazard ratios and confidence intervals assuming either the best or worst diet score for every individual, which only changed the population score range by 1.5 points. Using EHR data to identify outcomes has inherent limitations, including imperfect specificity of diagnostic codes. Some conditions may be miscoded, under-coded, or not captured if they are not recognized or documented during clinical care. We did not have access to individual notes to conduct chart review. Even so, our findings reflect how diagnoses are recorded in routine practice across diverse health systems, including large regional medical centers and federally qualified health centers. Finally, while the overall AoURP is highly diverse, the cohort we studied largely reflects the demographics of Fitbit consumers, which may not be generalizable.

## Conclusion

Higher LE8 scores were significantly associated with lower risk of CVD among Americans when using a combination of EHR and wearable device data. Our results emphasize the role of sleep health and physical activity in the calculation of LE8 scores and its association with incident CVD. Our findings suggest that using wearable device data linked to an EHR is a valuable, pragmatic, and effective method of monitoring population CVH and could pave the way for individualized CVH guidance over time, while minimizing limitations associated with patient-reported data.

## Data Availability

All data are publicly available at www.allofus.nih.gov following proposal submission and approval.

## Author Contributions

JOT, JA, ELB had full access to all of the data in the study and take responsibility for the integrity of the data and the accuracy of the data analysis. JOT, JA, ELB were responsible for concept and design. JA acquired, analyzed or interpreted data. JOT, JA, ELB drafted the manuscript. All authors performed critical revision of the manuscript for important intellectual content. JA did the statistical analysis. E.L.B. obtained funding. E.L.B. was responsible for supervision.

## Financial Disclosures

E.L.B has received research funds unrelated to this work from Anumana and unrestricted funds for research from Google.

Fitbit and Google were not involved in the collection, management, and analysis of the data, nor in the decision to submit the manuscript for publication.

The *All of Us* Research Program was not involved in the design and conduct of the study; collection, management, and analysis of the data; preparation, review, or approval of the manuscript; and decision to submit the manuscript for publication. To ensure privacy of participants, data used for this study was accessed and available to approved researchers only following registration, completion of ethics training, and attestation of a data use agreement through the All of Us Research Workbench platform, which can be accessed via https://workbench.researchallofus.org/login.

## DATA AVAILABILITY

To ensure privacy of participants, data used for this study are available to approved researchers following registration, completion of ethics training and attestation of a data use agreement through the All of Us Research Workbench platform, which can be accessed via https://workbench.researchallofus.org/.

## CODE AVAILABILTY

Code used for this study can be made immediately available to any approved researchers on the

*All of Us* Research Workbench platform by contacting our study team.

---

**Supplemental Table 1.**
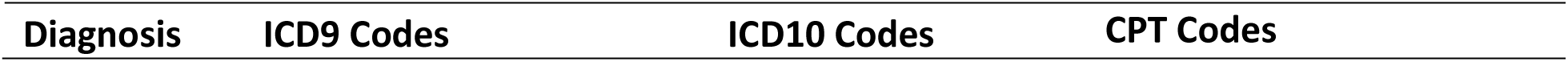
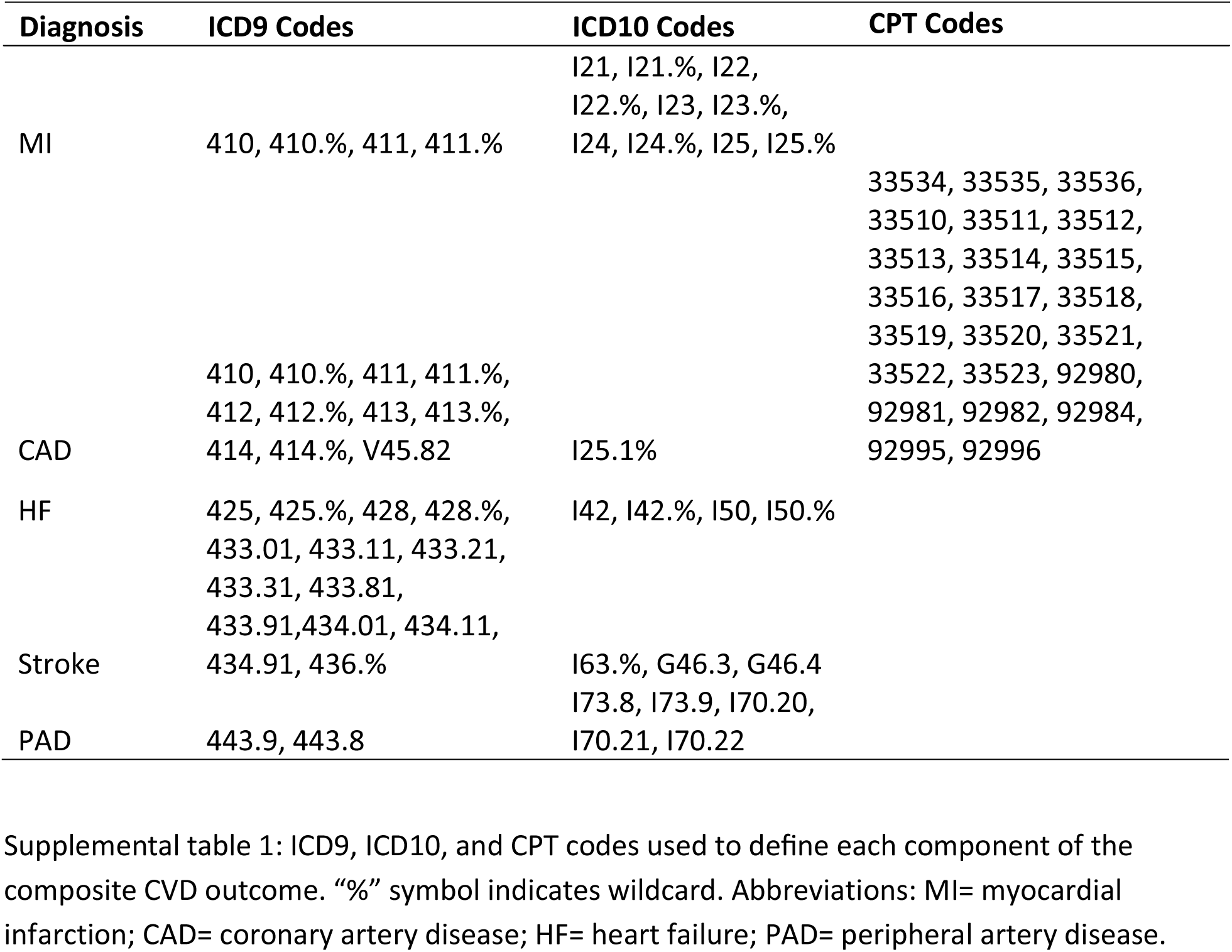
Definitions for each diagnosis.

**Supplemental Table 2.** : Sensitivity Analysis

Hazard ratios (HRs) and 95% confidence intervals (CIs) for incident CVD per 10-point increase in the LE8 total score, according to the maximum number of missing LE8 components permitted per participant (excluding diet).
| Number of missing components | Number of Events | HR (95% CI) |
| --- | --- | --- |
| ≤ 1 | 471 | 0.74 (0.67, 0.81) |
| ≤ 2 | 536 | 0.72 (0.66, 0.79) |
| ≤ 3 | 761 | 0.73 (0.67, 0.78) |

**Supplemental Table 3.**
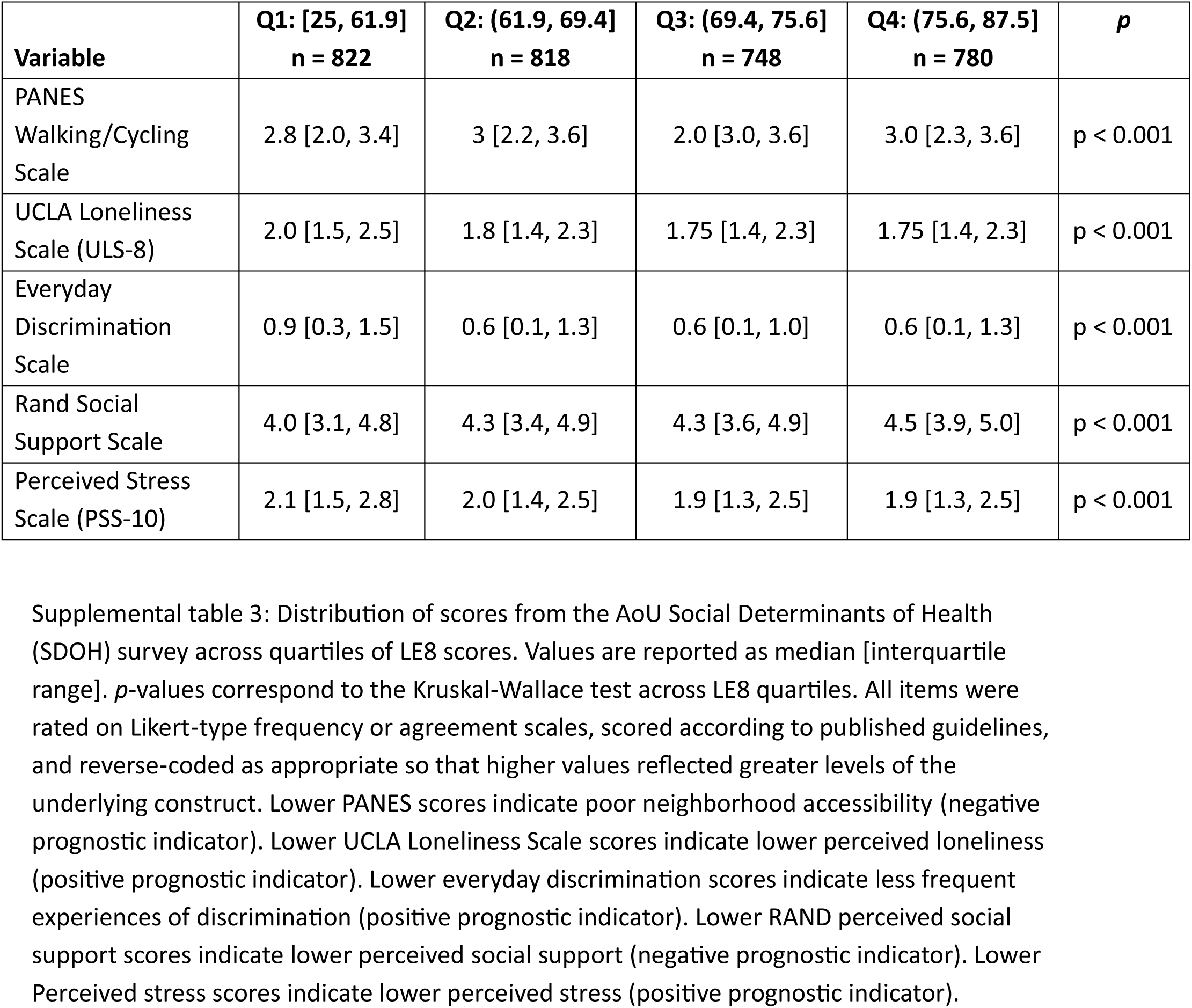
: Associations between LE8 Quartile and Psychological Stressors

Distribution of scores from the AoU Social Determinants of Health (SDOH) survey across quartiles of LE8 scores.
| **Variable** | **Q1: [25, 61.9]**<br>**n = 822** | **Q2: (61.9, 69.4]**<br>**n = 818** | **Q3: (69.4, 75.6]**<br>**n = 748** | **Q4: (75.6, 87.5]**<br>**n = 780** | ***p*** |
| --- | --- | --- | --- | --- | --- |
| PANES<br>Walking/Cycling<br>Scale | 2.8 [2.0, 3.4] | 3 [2.2, 3.6] | 2.0 [3.0, 3.6] | 3.0 [2.3, 3.6] | $p < 0.001$ |
| UCLA Loneliness<br>Scale (ULS-8) | 2.0 [1.5, 2.5] | 1.8 [1.4, 2.3] | 1.75 [1.4, 2.3] | 1.75 [1.4, 2.3] | $p < 0.001$ |
| Everyday<br>Discrimination<br>Scale | 0.9 [0.3, 1.5] | 0.6 [0.1, 1.3] | 0.6 [0.1, 1.0] | 0.6 [0.1, 1.3] | $p < 0.001$ |
| Rand Social<br>Support Scale | 4.0 [3.1, 4.8] | 4.3 [3.4, 4.9] | 4.3 [3.6, 4.9] | 4.5 [3.9, 5.0] | $p < 0.001$ |
| Perceived Stress<br>Scale (PSS-10) | 2.1 [1.5, 2.8] | 2.0 [1.4, 2.5] | 1.9 [1.3, 2.5] | 1.9 [1.3, 2.5] | $p < 0.001$ |

**Supplemental Figure 1.**
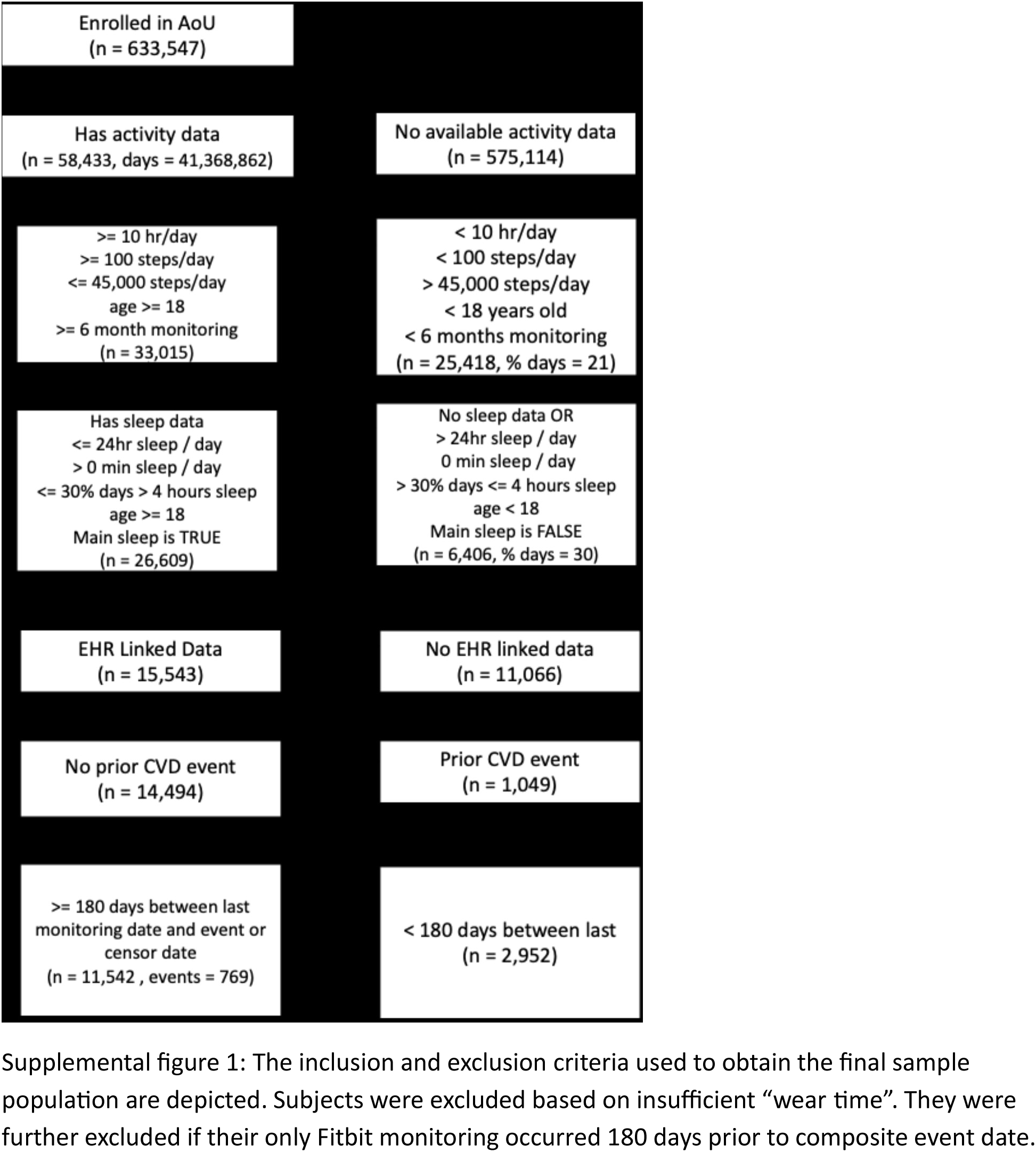
: Inclusion and Exclusion Flow Diagram

**Supplemental Figure 2a.**
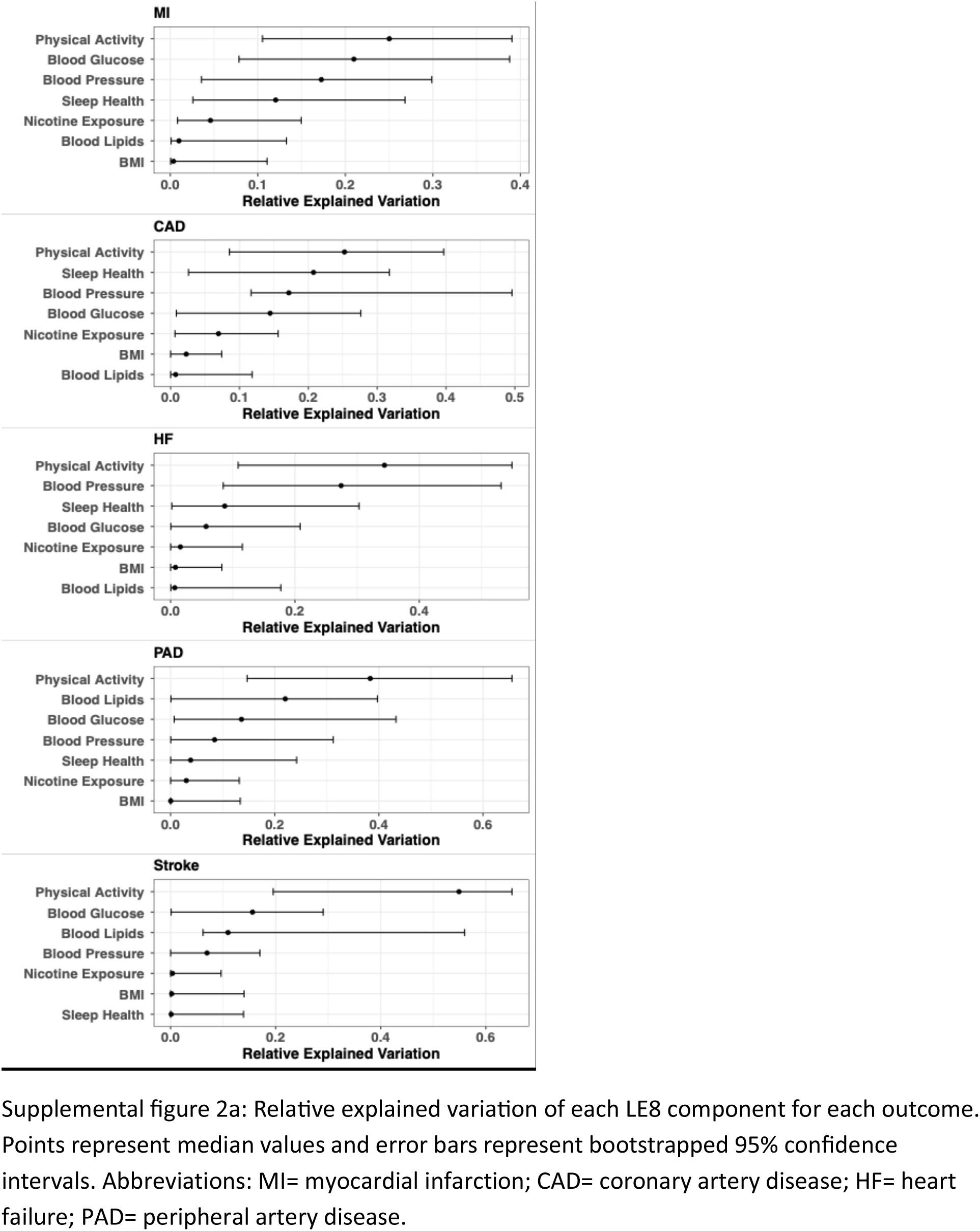
: Relative explained variation plot for each outcome.

**Supplemental Figure 2b.**
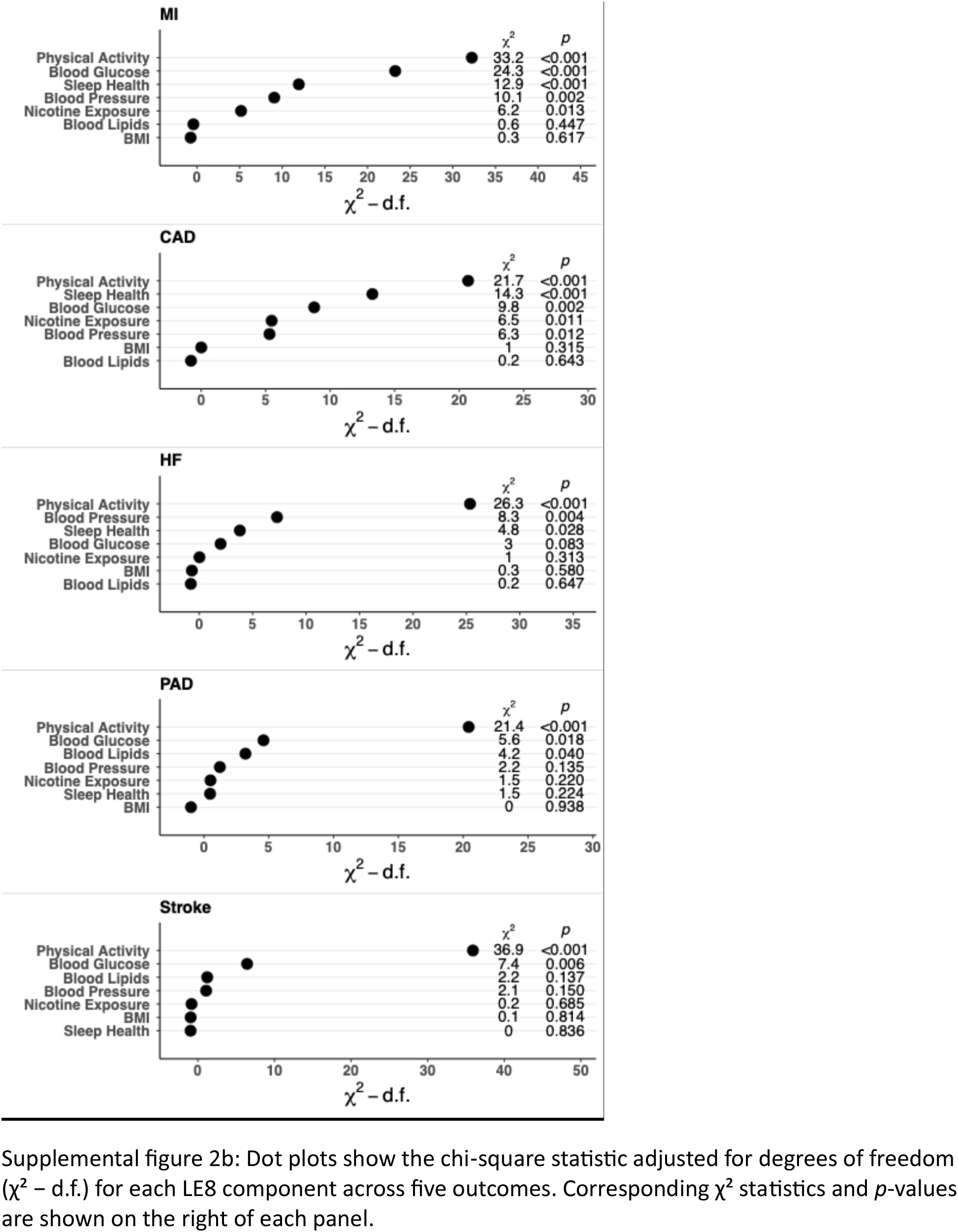
: Chi square table for the association of each LE8 component by outcome

**Supplemental Figure S3.**
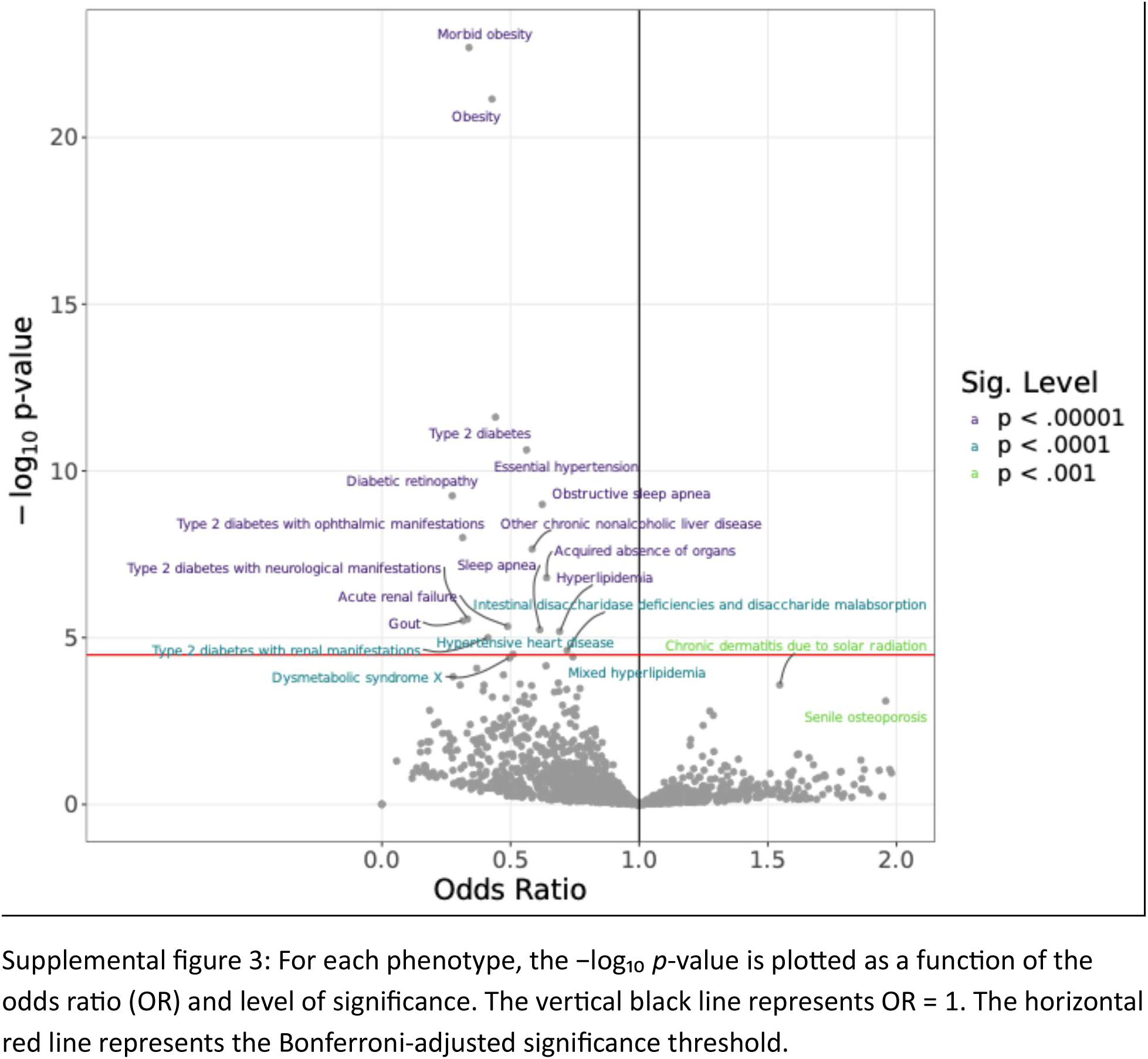
Volcano plot showing associations between clinical phenotypes and cardiovascular health.

